# Understanding the influence of leadership, organisation, and policy on delivering an integrated child health and social care service in community settings: A qualitative exploration using the SELFIE framework

**DOI:** 10.1101/2024.11.20.24317624

**Authors:** I Litchfield, L Harper, S Abbas, F Dutton, M Melyda, C Wolhuter, C Bird

**Affiliations:** Department of Applied Health Research, University of Birmingham, UK; Clinical Research Department, Primary Health Care Corporation, Qatar; Small Heath Medical Practice, Birmingham, UK; GreenSquareAccord, UK; Birmingham Women’s and Children’s NHS Foundation Trust, Birmingham, UK

**Keywords:** Integrated care, Health inequalities, Primary care, Children and young people, Community engagement

## Abstract

**Background:** The Sparkbrook Children’s Zone is an example of a place-based integrated health and social care service developed to support children and young people living in marginalized populations in the United Kingdom. This model of care is expected to address both clinical need and the social determinants of health but evidence of the practical support needed is lacking.

**Objective:** To understand the infrastructural challenges of providing a service combining clinical and non-clinical staff from a range of organisations and settings.

**Methods:** A qualitative exploration of the experiences of staff delivering the service and used a directed content analysis to present the results within the Sustainable integrated chronic care model for multi-morbidity: delivery, financing, and performance (SELFIE) framework.

**Results:** A total of 14 staff were interviewed including clinicians, social care providers, local voluntary groups, and school-based family mentors. Participants described the gap between system-level integration and the lack of practical support for delivering a unified service on the ground; the training opportunities afforded by collocation; the complexity of securing staff from multiple employers using various funding sources; and the need for lengthier evaluations that extend beyond early instability.

**Conclusions:** Despite decades of structural reform aimed at integrating the health and social care system in the UK, there was a surprising lack of practicable support for delivering a place-based integrated health and social care service. Their delivery is also hindered by short-term funding cycles limiting the reliability of evidence gathered from complex and evolving services.

**Research in Context:** *What is already known about the topic?:* Policymakers and commissioners in health systems worldwide are encouraging greater collaboration between health services, social care providers, and voluntary, community and faith sector groups to improve health outcomes and more effectively address the social determinants of health. Work on how precisely these integrated services might be configured is in its infancy and evidence of best practice is inconsistent.

*What does this study add to the literature?:* Participants described the gap between structural integration at system level and the lack of established process or infrastructure necessary to support a unified service on the ground. Those working in the service described how observational on-the-job training helped them understand the elements being delivered by different sectors. The complexity of negotiating with multiple employers and funding sources to secure staff was described, alongside the need for a lengthier period of evaluation that extends beyond short-term funding cycles.

*What are the policy implications?:* In the UK, policies for integrated care have resulted in the integration of high-level processes such as commissioning, strategic planning and financing. They now need to address practicable issues of infrastructure, targeted funding and administrative process necessary to support frontline provision of integrated care.

## Background

The increasing prevalence of chronic conditions, obesity, and mental ill health in high income countries is a particular concern for children, young people (CYP) and families belonging to minoritized, economically and culturally marginalized communities [1–3]. Their challenges are exacerbated by a range of socio-economic and cultural pressures that inhibit the utilisation of primary or preventative health care services [4–7]. Evidence suggests that integrated place-based health and social care can reduce health disparities, improve patient satisfaction, population health and cost effectiveness [8–10]. To this end policymakers and commissioners in multiple health systems worldwide are encouraging greater collaboration between health services, social care providers, local authorities, and voluntary, community and faith sector (VCFS) groups to improve health outcomes and more effectively address the social determinants of health (SDoH) such as income, housing, and food insecurity [11–18].

Work on how precisely these policy goals might best be achieved is in its infancy and evidence of best practice is inconsistent [19–24]. Meanwhile, in the UK, the latest attempts at reform have seen services restructured into integrated care systems combining primary, secondary, community and social care [25–27]. However delivering integrated health and social care at community level in the UK is challenging within a traditionally fragmented health and care system [28] with provider’s training and qualifications focused on a single speciality or setting [29–31], and a culture lacking in collaborative approaches to leadership or governance [32–36].

As attempts continue to overcome these barriers a number of pilots have emerged in the UK prioritizing localised delivery of preventative health and social care targeting CYP from underserved populations [8, 37, 38]. The Sparkbrook Children’s Zone (SCZ) is one such pilot service, collocating general practitioners, family support workers, mental health outreach, and paediatricians in a low-income area of Birmingham (UK) to deliver placed-based health and social care to underserved CYP (see Supplementary File 1 for a blueprint of the service) [39]. This challenging environment means it offers a valuable opportunity for an in-depth exploration of how these novel services perform and the contextual influences that impact their success. This paper uses qualitative data collected from staff delivering the service to populate an *a priori* framework developed to understand the broader contextual influences on delivering integrated care [40]. This has enabled the provision of structured insight into the processes and infrastructure that underpin the ability to deliver localised integrated care.

## Methods

### Study design

The work consists of a qualitative exploration of staff perspectives using data gathered from a series of semi-structured interviews and analysed using the “*Sustainable intEgrated chronic care modeLs for multi-morbidity: delivery, Financing, and performance”* (the SELFIE framework) [40]. The framework consists of a number of coordination concepts from micro-through to macro-levels incorporated within six domains informed by the World Health Organisation’s interpretation of healthcare systems (see Figure 1) [41]. The focus of this work was the surrounding contextual factors supporting delivery of the service and so the data was analysed using the domains of *Leadership & Governance, Workforce, Finance, and Information & Research* [40]. Our sister paper uses the SELFIE framework to describe the nature, and content of the service being delivered [42].

**Figure 1.**
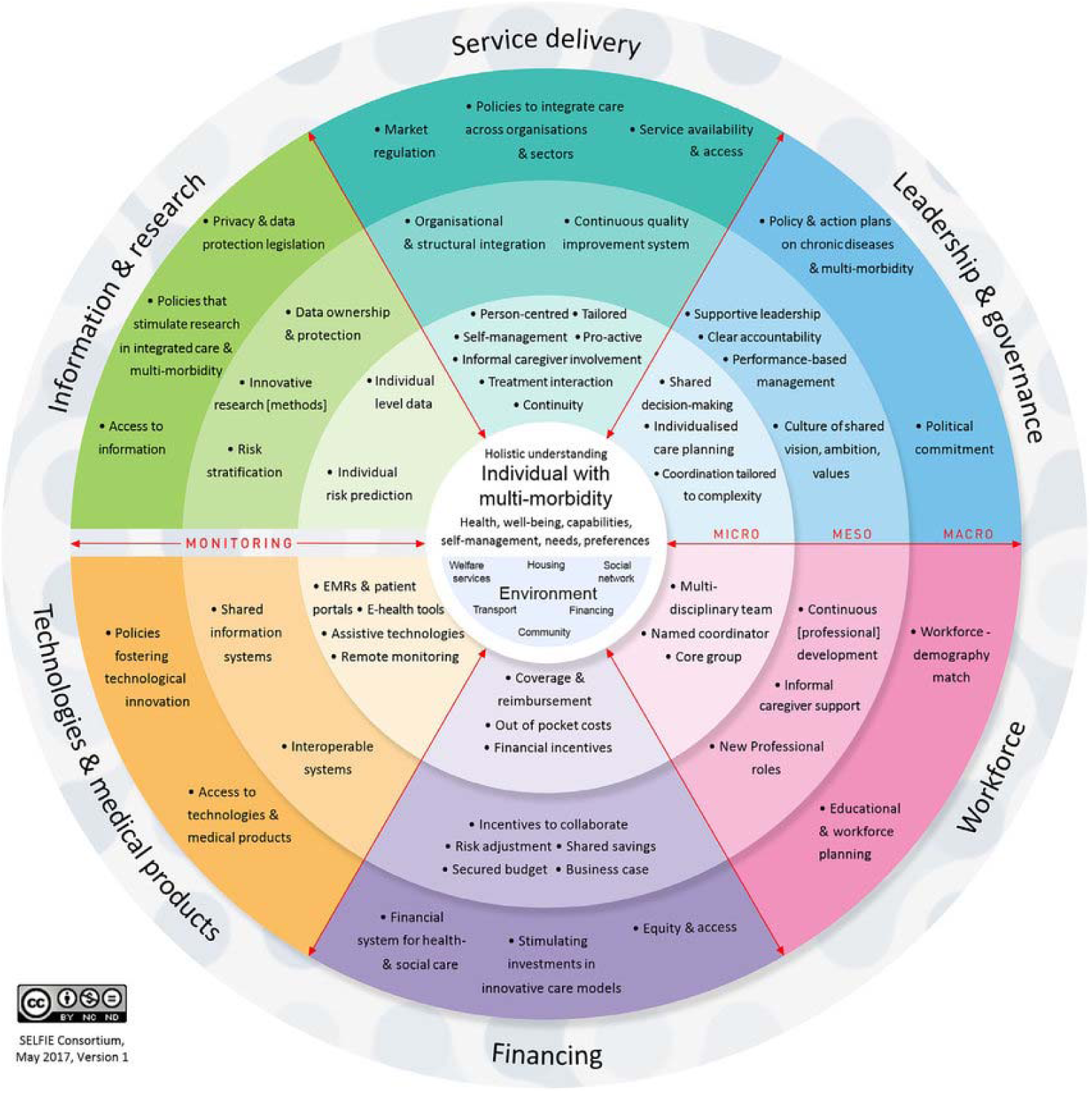
The SELFIE framework. [40]

### Population/recruitment

The SCZ is based in the Sparkbrook & Balsall Heath East ward in Birmingham, a large and diverse city in the UK’s midlands. It is the second most populous ward in the city, has the second highest level of deprivation and a superdiverse, young population with high rates of unemployment and one of the highest levels of infant mortality in England [43]. It is also disproportionately affected by childhood obesity, child criminal and sexual exploitation, poor housing, chronic disease, and high levels of universal needs around housing, food, clothing, sanitary products, and essential supplies [43].

All staff involved in developing, managing and delivering the SCZ were eligible for inclusion. They were approached by [1st author] and [7^th^ author] via email and in-person all were supplied with a participant information sheet, and the opportunity to ask questions of their participation; ultimately providing informed consent before the interview commenced. We aimed to carry out interviews with 5-6 service providers from each organisation (including service leads, those actively delivering the service and administrative/support staff) to reach a total of 25 interviews sufficient to provide a rich and representative data set [44].

### Data Collection

Semi-structured interviews were conducted online (via Teams or Zoom), face-to-face in a room at the clinic, or via telephone by [First author] and [Third author]. They are experienced qualitative researchers, unknown to participants, that used a topic guide informed by the existing literature and with questions and prompts relating to a range of themes including experiences of engaging with the local Integrated Care System, barriers and facilitators to delivering the SCZ, and reflections on its future development. Digital audio recordings were transcribed verbatim by an approved third-party transcription service and the data were managed using nVivo vs12.

### Data analysis

Two authors [first author] and [third author], experienced qualitative researchers, independently coded each transcript fitting the data within each of the relevant themes of the SELFIE framework using a directed content analysis [45] that allowed the identification and inclusion of emerging domains, constructs or sub-constructs [46]. Any differences in coding were discussed between the two authors and a consensus arrived at. The final allocation of the data within the coding framework was agreed by all authors.

## Results

### Characteristics of participants

We interviewed 14 participants over 13 interviews (two participants were interviewed at the same time). The interviews lasted between 18 and 70 minutes. Of the 14 participants five were from primary care, three secondary care, two from social support, one that worked in local education, and one for a children’s charity.

### Qualitative results

Our emergent findings are presented below within each of the four named domains and pre-determined constructs of the SELFIE framework. They are described alongside exemplar quotes identified by Participant ID, Sector, and Job role. These findings are summarised in Table 2.

**Table 1.** Characteristics of participants

| ***Participant ID*** | ***Sector*** | ***Role/Job Title*** |
| --- | --- | --- |
| 01 | Secondary care | Senior staff |
| 02 | Education | Family mentor |
| 03 | Primary Care | GP |
| 04 | Secondary Care | Consultant |
| 05 | Secondary Care | Consultant |
| 06 | Primary Care | GP |
| 07 | Social support | Family Support |
| 08 | Integrated care | Operations manager |
| 09 | Primary care | Health Care |
| 10 | Primary care | Health Care |
| 11 | Children's charity | Service lead |
| 12 | Primary care | GP |
| 13 | Social support | Project manager |
| 14 | Social support | Service lead |

**Table 2.** Summary of SELFIE informed analytical framework and emerging themes [40]

| **Domain** | *Definition* | *Level* | **Construct** | **Example from SCZ** |
| --- | --- | --- | --- | --- |
| **1. Leadership & Governance** | The strategic policy frameworks and organisational oversight that shape the delivery of care. Includes regulation, system design, and accountability. | *Micro* | Coordination tailored to complexity | The provision of collocated health and social care/support |
|  |  | *Meso* | Shared vision/decision making | Open lines of communication with senior staff (shared learning), Joint purpose |
|  |  | *Macro* | Political commitment | Lack of integrated infrastructure, |
|  |  |  | Policy and action plans | Community engagement, Organic development |
| **2. Workforce** | The knowledge, skills, motivation and deployment of the people responsible for organizing and delivering health and social care services. | *Micro* | Multi-Disciplinary Team | Seamless referrals between agencies |
|  |  | *Meso* | Continuous development | Training in preventative care, improved understanding of other elements of service |
|  |  | *Macro* | Demography match | Multi-lingual workforce |
|  |  |  | Educational and workforce planning | Need for administrative infrastructure and support, Opportunities for non-clinical roles |
| **3. Financing** | The allocation of money to cover the health needs of the people, individually and collectively, includes both core funding and service incentives. | *Micro* | Coverage and reimbursement | Different budgets and employers |
|  |  | *Macro* | Stimulating investment in innovative services | Perceptions of duplication, Excessive bureaucracy |
|  |  |  | Finance system for health and social care | System wide lack of resource |
| **4. Information & Research** | The health-related information generated that can facilitate analysis and synthesis and underpin evidence-based health and social care. | *Macro* | Access to information | Appropriate time scale, Evaluation of process as well as outcome |

## 1. Leadership & Governance

### Coordination tailored to complexity (<u>Micro</u>)

Participants described how the SCZ was designed to treat the patient and address the SDoH within a single place-based service, and demonstrated a shared understanding that this was inherently linked to the combination of clinical care and social support:

> “…reducing the inappropriate use of A&E, is the NHS ambition, and that’s their thing… in Early Help our objective is to prevent families from going into crisis, and we can’t do that on our own, so we can’t do that just in the community sector or the voluntary sector, we have to have doctors and GPs, dentists and youth workers…to come in and influence how a family build resilience. So SCZ is a mechanism of doing that…we can do our best to support a person who has severe asthma by improving their home situation, so reducing mould, getting better living conditions, getting them to be more active, get them outside of the house more, but if they don’t have the inhaler, or they don’t have the opportunity to go and see a doctor?…our objective is really to be working together…[as] it’s never a single issue…”
>
> P013, Social Support, Project manager

### Shared vision/decision making (<u>Meso-)</u>

Staff reflected on how service leads were clear on what the aims of the service were and how they should be met. This included the fostering of an open learning culture within the organisation where senior service leads were equally happy to offer advice as to welcome challenge and enquiry:

> “…the people who are working at the Children’s Zone they’re all just very open. There’s no ‘boundaries’, there’s no ‘silly questions’… and shared learning just flourishes really…’we’ll learn from you, and you’ll learn from us’ and that’s what’s happened.”
>
> P12, Primary Care, GP

### Political commitment (<u>Macro-)</u>

Despite a regional and national drive towards integration that has lasted decades [47], participants described the lack of any *a priori* processes or underpinning logistical infrastructure necessary to deliver a combined place-based service:

> “I think there’s an awful lot of talk about integration, and actually there’s very little experience of it having been done, and of the challenges then that go with it, and that’s all I talked about with the logistics, that we don’t have shared emails, and that the governance how do you define who does that? Who gets a CQC registration when it’s a joint clinic? It’s so many things like that, that can be really, really challenging, and I don’t think the big organisation of the NHS is in any way geared up really to managing that level of complexity.”
>
> P04, Secondary Care, Consultant

### Policy and action plans (<u>Macro-)</u>

Participants described how the creation of the SCZ evolved from following children repeatedly presenting at the local emergency department that could have been treated in primary care was mistakenly sent to the emergency department. In the early stages of development that followed, the leads of the SCZ recognised the importance of understanding the preferences of the community and the need to build trust in mainstream healthcare [48]. This involved a period of consultation lasting several months and enabled the service that emerged to better reflect local socio-cultural sensitivities:

> “I know it did take time for us to build trust in that community - so it can’t be like a roving thing that you land somewhere and then move somewhere in three months - it takes three months for that trust to build, and it takes a while for that to embed in professional networks but also community networks. So, once you’ve got that then you can keep going. So, it’s structuring KPIs, and whatever other measures they’ll put in to allow for that community engagement, to allow to that trust to be built from professionals and the community…”
>
> P11, Children’s Charity, Service Lead

## 2. Workforce

### Multi-Disciplinary Team (<u>Micro-)</u>

The SCZ was predicated on creating a multi-disciplinary team that not only provided the key tenets of health and social care and support but also had established links into other services and community groups such as those that provided well-being counselling or preventative care:

> “…the majority of time I really do feel fulfilled going there. I do feel like I have the opportunity to point out gently and kindly [to families] the areas where behavioural change does need to happen for various lifestyle factors like obesity, diet, parenting, whatever…and I crucially got the support to be able to do it.”

P04, Secondary Care, Consultant

### Continuous development (<u>Meso-)</u>

Many of the clinicians involved in delivering the SCZ were required to develop additional skill sets and develop a greater understanding of how other sectors worked. For example, the lead of the health and well-being counselling service (“PAUSE” [49]) that was part of the SCZ explained how they would invite clinicians to observe how they go about their work:

> “…one of the key things for us when we start working in partnership with anybody is to get that practitioner - whatever their inexperience - into the room to see how it works. Because it’s very different, and they need to be able to prepare that parent, or carer, or young person for that.”
>
> P11, Children’s Charity, Service Lead

### Demography match (<u>Macro-)</u>

There was an awareness of the super-diverse population the SCZ was serving and the importance of providers being able to communicate in languages which reflected that cultural diversity:

> “We have to adapt our communication style… a number of our team are able to speak multiple different languages, so in our team we can speak Punjabi, Urdu, Arabic, Farsi, French, there’s quite a few other languages, and that really enables us to offer a service that feeds into the needs of our families.”
>
> P13, Social Support, Project Manager

### Staff education and planning

Senior service leads lamented the lack of the administrative support required to underpin an integrated service with much of the burden falling to clinical staff. For example, this included the absence of a dedicated project manager that could have alleviated some of the managerial burden placed on clinicians. Another specific example provided was the necessity for dedicated human resource support to manage the complexities of ensuring even a single clinician could work in the SCZ clinic once a week:

> “…the other thing about the local integration - which I think has to be acknowledged - is that if you’re having a doctor coming every week, to come and do the work, there is an impact on whoever is doing HR. I know this sounds really small, but again it’s one of those things that took us by surprise… there has to be quite a process to make sure that person has adequate DBS checks, that they’ve got the appropriate indemnity in place, that they are registered and you have their band details…for one session a week service it’s been quite admin heavy!”
>
> P08, Integrated care system, Operations Manager

## 3. Finance

### Coverage and reimbursement (<u>Micro-</u>)

Securing and paying for staff drawn from multiple organisations and funding bodies presented difficulties in the absence of a pre-defined process to secure funds or enable payment across settings:

> “…we’ve had a bit of a challenge … for instance, our eczema support was only available on a trial period…dental [care] we were really keen to get on-board and that hasn’t quite worked out. There’s been just some teething problems around how people are employed and paid, because this is an integrated model and we’re all working in different places, and none of us have a unified employer.”
>
> P05, Secondary care, Consultant

### Finance system for health and social care (<u>Macro-)</u>

The SCZ has been developed and delivered within a health and social care system that has been chronically underfunded for over a decade [50]. Reflective of this broader lack of resources was the limitations on the amount of (paid) time clinicians were able to devote to the SCZ.

> “…where I used to work they would have hired someone to do this [delivery of the SCZ] as their full time job, but instead you’ve got [consultant] working in A&E, [GP] doing her GP plus A&E and you just think… I don’t know, in your free time…? you’re then trying to do this? So yeah, I would say that is a big problem…there’s just not enough resource.”
>
> P13, Social Support, Project Manager

### Stimulating investment (<u>Macro-)</u>

The SCZ was purposely designed to gather multiple services together in a single location which risked the perception described by one senior service lead that it was being funded to repeat work that individual organisations already provided elsewhere, or otherwise “needs to move more towards using non-doctors” (P03, GP) to bring costs down. Meanwhile, the inhibitive nature of the bureaucracy that surrounded establishing innovative services such as the SCZ was described:

> “I wish there could be an element of cutting through stuff, and that can happen across the board really, cutting through the red tape to get somewhere to be like ‘…can you just do it! You’ve got a good idea…’ it can really suck the energy out of everyone in the room, the requirements to go through to process - all this ‘stuff’. I don’t know, I think there is a way to do things safely, which doesn’t have to stifle everything.
>
> P04, Secondary Care, Consultant

## 4. Information and research

### Access to information (<u>Macro-)</u>

There was a broad understanding amongst those delivering the service of the need to capture appropriate data to demonstrate the worth of the SCZ to patients and the health and social care system. There was also agreement that any evaluation should not focus solely on outcome but also explore the process and methods of working and how they might be replicated at scale. Because of the novelty of the service participants also felt that data should be collected over a longer time scale, allowing the service to reach a degree of maturity and stability:

> “…it takes time to embed the service or clinicians to get used to changing their patterns of referral patterns and things, and having confidence and trust in the new service…So that’s why I say it needs more time to be evaluated and given time to come to fruition.”
>
> P06, Primary Care, GP

## Discussion

### Summary of findings

The use of the SELFIE framework enabled structured insight into the complex influences informing the delivery of place-based integrated health and social care. Related to the domain of *Leadership & Governance* participants described the importance of supportive and accessible leadership of the service though noted the gap between structural integration at system level and the lack of process and infrastructure necessary to support a unified service in real-world environments; Regards implications for the *Workforce*, staff described how on-the-job training helped them understand all elements of the service; In relation to *Finance* the complexity of negotiating with multiple employers and funding sources to secure staff was described, alongside suggestions for how investment in integrated services might be encouraged; finally, in relation to *Information and research,* the need for a lengthier period of evaluation that extends beyond the instability of the early phases of implementation to include more reliable data was described.

### Strengths and Limitations

Our rich dataset has provided valuable data on some of the contextual influences impacting the delivery of one of the UK’s first collocated place-based, integrated health and social care services. Participants were representative of the organisations involved in delivering the service and though their number (n=14) was lower than anticipated, the majority of active staff were interviewed, highlighting the issues in recruiting and funding staff in the early phases of the SCZ. The SELFIE framework proved a valuable tool in unpicking the experiences of delivering a collocated cross-sector community-based service and we used best practice in directed content analysis without constraining the results [45, 46]. The validity of the findings was supported by regularly sharing and discussing the outputs of the analysis across the team [51]. Not every element of the SELFIE was identified in our data set though the comprehensive nature of the framework meant that it accommodated all of our data.

### Specific findings

#### Leadership & Governance

The latest research across multiple industries recommends diminished, hierarchical distinctions between managers and employees [52–54]. However, in healthcare the structure remains typically hierarchical, with power focused on a handful of groups that risks compromising teamwork and patient safety [55–57]. In contrast we heard how senior SCZ leads created a work culture that promoted shared learning and decision making which as previously, was associated with increased job satisfaction and greater mutual respect across disciplines [58, 59]. Similarly, the collective leadership demonstrated by senior SCZ leads, characterised by their shared goals and responsibilities, has been linked with more sustainable collaborative relationships [60–63].

In the UK, the broad policy goals of better integrated health and social care have existed for years [64]. Yet to date these have tended to focus on integrating high-level processes such as commissioning, strategic planning and financing and the creation of joint boards, forums and committees [16, 64, 65]. What became apparent within our work was the lack of supporting infrastructure, and administrative process necessary to support front-line integration. In other health systems such as in the United States standardized care coordination protocols are common across those programs that bridge health care and social services [66]. The absence of similar in the UK may in part be due to the variable and *ad hoc* nature of many localised integrated care offers that has precluded generation of shared learning and development of similar national protocols [20]. At least within the UK, it appears that it is now time for policymakers to reflect on the practicalities of delivering integrated care at local level and how existing systems might be adapted in their support [67, 68].

One important element in establishing integrated services that is more widely recognised is the early involvement of local communities in the design of the service, particularly those most vulnerable or seldom heard [69]. Senior leads at the SCZ undertook and learnt from an extensive period of community consultation and such “meaningful, and trusted” engagement. This engagement is stipulated in the statutory guidance for Integrated Care Boards and otherwise considered best-practice in developing place-based partnerships [70, 71] and ensuring services are context-specific [72], sympathetic to community cultural and social norms [73],

#### Workforce

Staff described the value of observing consultations delivered by colleagues from different sectors, such as the children’s counselling service [49]. Similar observational training techniques have been employed in other healthcare environments where they have helped build competencies in inter-professional collaborative practice [33, 74–76], and improved understanding and respect for healthcare professionals from other sectors [33, 77, 78]. Specifically in support of integrated care in the UK, the NHSE has developed a number of training initiatives to support cross-discipline collaboration [25, 35, 79, 80], including training in collaborative leadership [32, 81].

It was reported how clinicians donated their own time to meet the needs of the responsibilities to the SCZ, reflective of broader workforce shortages in the NHS [82]. The need for cost-effective alternatives to support busy clinicians was voiced, and the NHS has introduced the Additional Roles Reimbursement Scheme (ARRS) to develop non-clinical roles, such as social prescriber or care navigator in primary care which have the potential to usefully augment an integrated care service such as the SCZ [83–85].

In relation to non-clinical roles, participants described the need for dedicated administrative support, recognised as a key element in successfully delivering integrated care [86, 87]. Not only can patient facing roles support access [88, 89] but the importance of back-office support was underestimated by the leads at SCZ, particularly in the absence of mechanisms to readily manage staff drawn from multiple organisations [66, 87]. Instead, many of these administrative duties became another demand on overly burdened clinical staff, likely inhibiting the efficiency of the service [90, 91].

#### Finance

Over the last decade funding has decreased across the NHS, social care, and public health [50, 92, 93], and yet integrated services are expected to provide not only high quality health care but also effectively addresses SDoH [94]. In this financially constrained environment it has been recommended that funding models are created that share financial risk, reward, and accountability across health and social care sectors [95–99], moving away from existing single-condition, target driven incentives, that currently predominate [100, 101]. Ideally this funding in health and social care would be allied with investment in broader social initiatives to improve housing, or food security [94, 102–104].

Despite the high level recommendations and priorities placed on integrated care, SCZ found itself competing for future funds with other services within their local ICS and impacted by a lack of funding for social care and unexpected cuts to integrated care board running costs [105]. Where investment in integrated care has been committed over the longer term the benefits of collaborative, place-based programmes such as the UK’s Sure Start are seen and realised [106].

#### Information and research

Iterative cycles of organisational and structural reform have failed to stimulate the desired routine integration of services on the front line due in part to a lack of evidence demonstrating either clear benefit or best practice [20]. Participants understood the importance for robust data collection and evaluation, though were aware of the tensions between the SCZ’s aim of long-term improvement and the short-term context of its implementation and opportunity to show benefit [107–109]. The need to demonstrate early and clear benefit is not unique to integrated services but the complexity of their offer means they require longer to become embedded than many other interventions [110–112].

Demonstrating benefit is also hindered by the difficulties in collectively measuring a range of interacting outcomes across clinical care, preventative care, and social care and support and compounded by a lack of comparable data against which to assess change [113]. Perhaps because of this, and the exaggerated influence of secondary care trusts, previous attempts at evaluating the impact of integrated care have been over reliant on hospital-based measures failing to capture the impact on health and social care [114]. It’s important that those funding integrated care services understand where and how the mechanisms of this integration create improved outcomes and the timescales within which they might be observed [113, 114].

## Conclusions

Despite closer integration being considered as the future for health and social care, the ability to deliver place-based integrated care was inhibited by uncertainty around dedicated funding, and a lack of supporting process and infrastructure. However, there are positive elements that emerged from the SCZ that could be usefully adopted more broadly, including observational training, flattened hierarchies, meaningful community engagement, and an open collaborative culture.

## Data Availability

All data produced in the study are available upon reasonable request to the authors

